# Rapid Detection of 2019 Novel Coronavirus SARS-CoV-2 Using a CRISPR-based DETECTR Lateral Flow Assay

**DOI:** 10.1101/2020.03.06.20032334

**Authors:** James P. Broughton, Xianding Deng, Guixia Yu, Clare L. Fasching, Jasmeet Singh, Jessica Streithorst, Andrea Granados, Alicia Sotomayor-Gonzalez, Kelsey Zorn, Allan Gopez, Elaine Hsu, Wei Gu, Steve Miller, Chao-Yang Pan, Hugo Guevara, Debra A. Wadford, Janice S. Chen, Charles Y. Chiu

## Abstract

An outbreak of novel betacoronavirus, SARS-CoV-2 (formerly named 2019-nCoV), began in Wuhan, China in December 2019 and the COVID-19 disease associated with infection has since spread rapidly to multiple countries. Here we report the development of SARS-CoV-2 DETECTR, a rapid (∼30 min), low-cost, and accurate CRISPR-Cas12 based lateral flow assay for detection of SARS-CoV-2 from respiratory swab RNA extracts. We validated this method using contrived reference samples and clinical samples from infected US patients and demonstrated comparable performance to the US CDC SARS-CoV-2 real-time RT-PCR assay.

---

Over the past 40 years, there have been recurrent large-scale epidemics from novel emerging viruses, including human immunodeficiency virus (HIV), SARS and MERS coronaviruses, 2009 pandemic influenza H1N1 virus, Ebola virus (EBOV), Zika virus (ZIKV), and most recently SARS-CoV-2^1,2^. All of these epidemics presumably resulted from an initial zoonotic animal-to-human transmission event, with either clinically apparent or occult spread into vulnerable human populations. Each time, a lack of rapid, accessible, and accurate molecular diagnostic testing has hindered the public health response to the emerging viral threat.

In early January 2020, a cluster of cases of pneumonia from a novel coronavirus, SARS-CoV-2 (with the disease referred to as COVID-19), was reported in Wuhan, China^1,2^. This outbreak has spread rapidly, with over 90,000 reported cases and 3,000 deaths as of March 4th, 2020^3^. Person-to-person transmission from infected individuals with no or mild symptoms has been reported^4,5^. Assays using quantitative reverse transcription-polymerase chain reaction (qRT-PCR) approaches for detection of the virus in 4-6 hours have been developed by several laboratories, including an Emergency Use Authorization (EUA)-approved assay developed by the US CDC^6^. However, the typical turnaround time for screening and diagnosing patients with suspected SARS-CoV-2 has been >24 hours given the need to ship samples overnight to reference laboratories. To accelerate clinical diagnostic testing for COVID-19 in the United States, the FDA on February 28th, 2020 permitted individual clinically licensed laboratories to report the results of in-house developed SARS-CoV-2 diagnostic assays while awaiting results of an EUA submission for approval^7^.

Here we report the development and initial validation of a CRISPR (clustered regularly interspaced short palindromic repeats)-Cas12 based assay^8-11^ for detection of SARS-CoV-2 from extracted patient sample RNA in ∼30 min, called SARS-CoV-2 DETECTR. This assay performs simultaneous reverse transcription and isothermal amplification using loop-mediated amplification (RT-LAMP)^12^ from RNA extracted from nasopharyngeal or oropharyngeal swabs in universal transport media (UTM), followed by Cas12 detection of predefined coronavirus sequences, after which cleavage of a reporter molecule confirms detection of the virus. We first designed primers targeting the E (envelope) and N (nucleoprotein) genes of SARS-CoV-2 (**Fig. 1a**). The primers amplify regions that overlap the WHO assay (E gene region) and US CDC assay (N2 region in the N gene)^6,13^, but are modified to meet design requirements for LAMP. We did not target the N1 and N3 regions used by the US CDC assay, as these regions lacked suitable protospacer adjacent motif (PAM) sites for the Cas12 guide RNAs (gRNAs). Next, we designed Cas12 gRNAs to detect three SARS-like coronaviruses (SARS-CoV-2 accession NC_045512, bat SARS-like coronavirus (bat-SL-CoVZC45, accession MG772933), and SARS-CoV, accession NC_004718)) in the E gene and specifically detect SARS-CoV-2 only in the N gene **(Supplementary Fig. 1)**.

**Figure 1.**
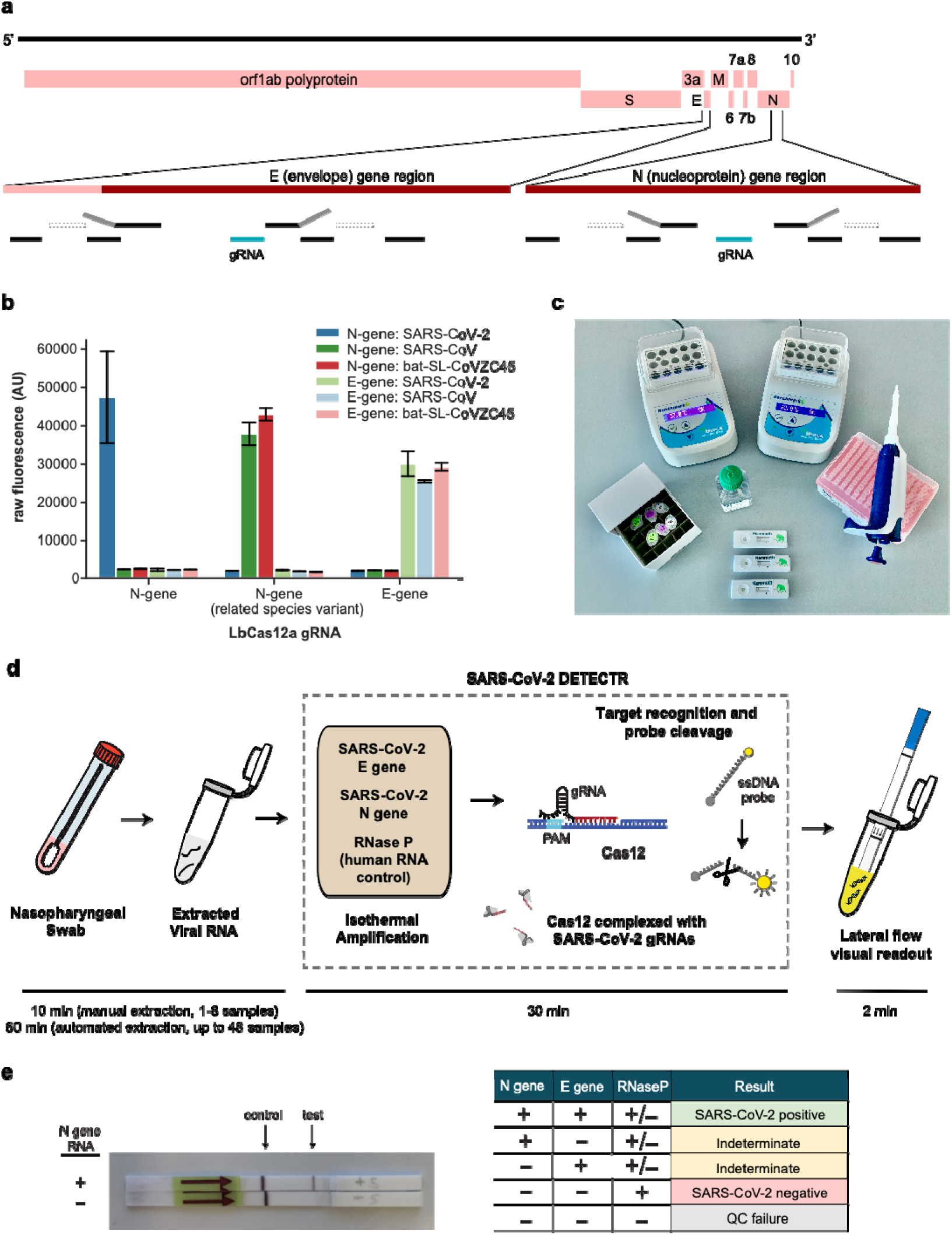
A CRISPR-Cas12 based assay for detection of SARS-CoV-2. **(a) Genome map showing primers, probes and gRNAs**. Visualization of primers and probes on the SARS-CoV-2 genome **(b) gRNA specificity**. Cas12 gRNAs are programmed to specifically target SARS-CoV-2 or broadly detect related coronavirus strains. The N gene gRNA used in the assay (left) is specific for SARS-CoV-2, whereas the E gene gRNA is able to detect 3 SARS-like coronavirus (right). A separate N gene gRNA targeting SARS-CoV and a bat coronavirus and differing by a single nucleotide from the N gene gRNA used in the assay fails to detect SARS-CoV-2 (middle). **(c) Minimum equipment needed to run protocol**. With appropriate biosafety level 2 requirements, the minimum equipment required to run the protocol includes Eppendorf tubes with reagents, heat blocks or water bath (37°C and 62°C), nuclease-free water, pipettes and tips, and lateral flow strips. **(d) Schematic of SARS-CoV-2 DETECTR workflow**.Conventional RNA extraction or sample matrix can be used as an input to DETECTR (LAMP pre-amplification and Cas12-based detection for E gene, N gene and RNase P), which is visualized by a fluorescent reader or lateral flow strip. **(e) Lateral flow strip assay readout**. A positive result requires detection of at least the two SARS-CoV-2 viral gene targets (N gene and E gene).

Using synthetic, *in vitro* transcribed (IVT) SARS-CoV-2 RNA gene targets in nuclease-free water, we demonstrated that the CRISPR-Cas12 based detection can distinguish SARS-CoV-2 with no cross-reactivity for related coronavirus strains **(Fig. 1b, Supplementary Fig. 2)**. We then optimized the conditions for the SARS-CoV-2 DETECTR assay on the E gene, N gene and the human RNase P gene as a control, which consists of an RT-LAMP reaction at 62°C for 20 min and Cas12 detection reaction at 37°C for 10 min. The DETECTR assay can be run in approximately 30 min and visualized on a lateral flow strip **(Fig. 1c, d)**. The SARS-CoV-2 DETECTR assay requires detection of both the E and N genes to confirm a positive test **(Fig. 1e)**, and interpretation is consistent with that for the CDC assay N1 and N2 genes (the N3 gene target region for the CDC assay is no longer being used due to concerns regarding flaws in manufacturing reagents and potential decreased sensitivity)^14^.

We next compared the analytic limits of detection (LoD) of the RT-LAMP/Cas12 DETECTR assay relative to the US FDA Emergency Use Authorization (EUA)-approved CDC assay for detection of SARS-CoV-2 **(Table 1; Fig. 2d)**. A standard curve for quantitation was constructed using 7 dilutions of a control IVT viral nucleoprotein RNA (“CDC VTC nCoV Transcript”)^6^, with 3 replicates at each dilution (**Fig. 2d, left; Extended Data 1)**. Ten two-fold serial dilutions of the same control nucleoprotein RNA were then used to run the DETECTR assay, with 6 replicates at each dilution (**Fig. 2d, right; Supplementary Fig. 3)**. The estimated LoD for the CDC assay tested by California Department of Public Health was 1 copy/µL reaction, consistent with the analytic performance in the FDA package insert, versus 10 copies/µL reaction for the DETECTR assay.

**Table 1.**
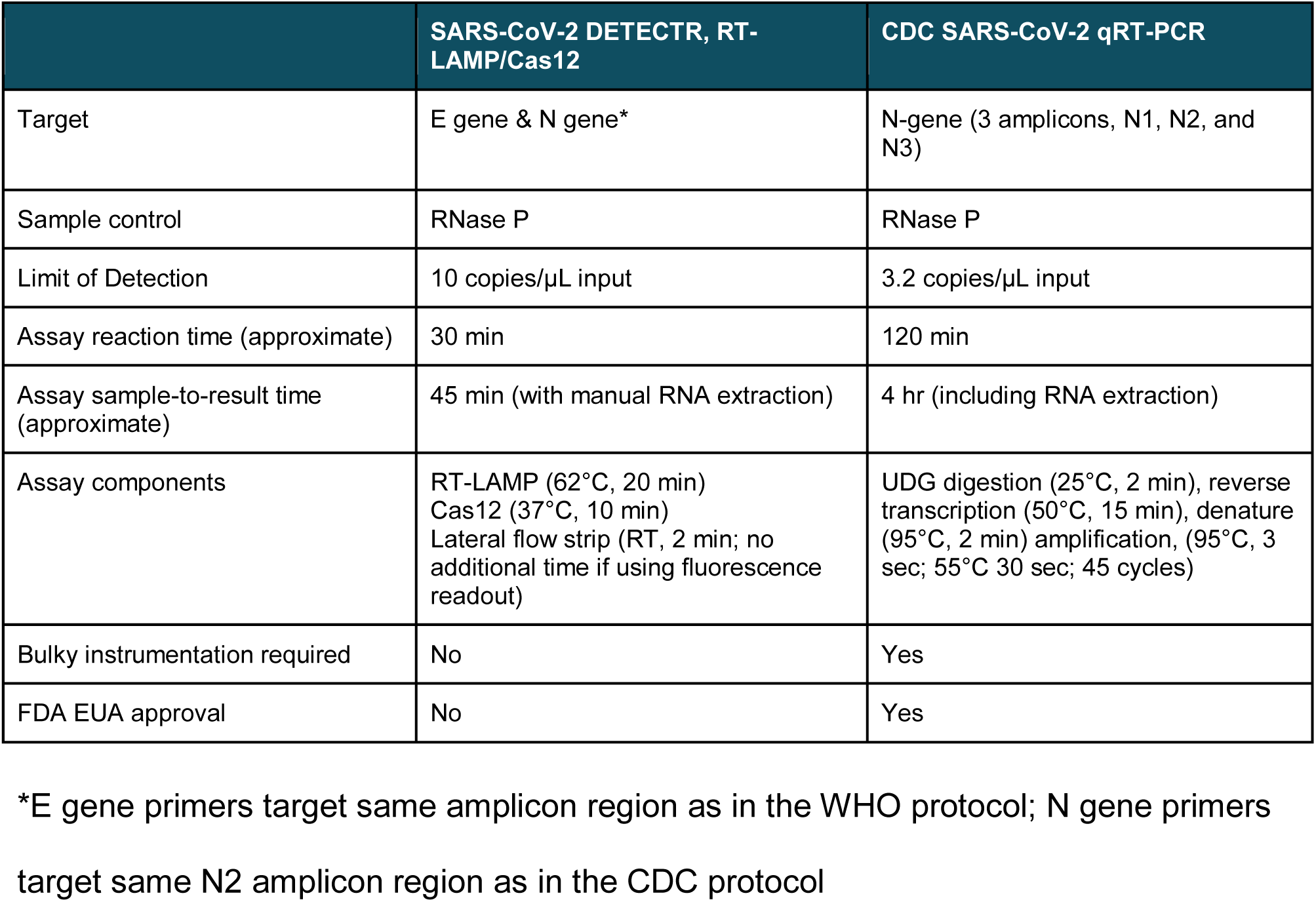
Comparison of the DETECTR (RT-LAMP/Cas12) assay with the CDC qRT-PCR assay for detection of SARS-CoV-2.

**Figure 2.**
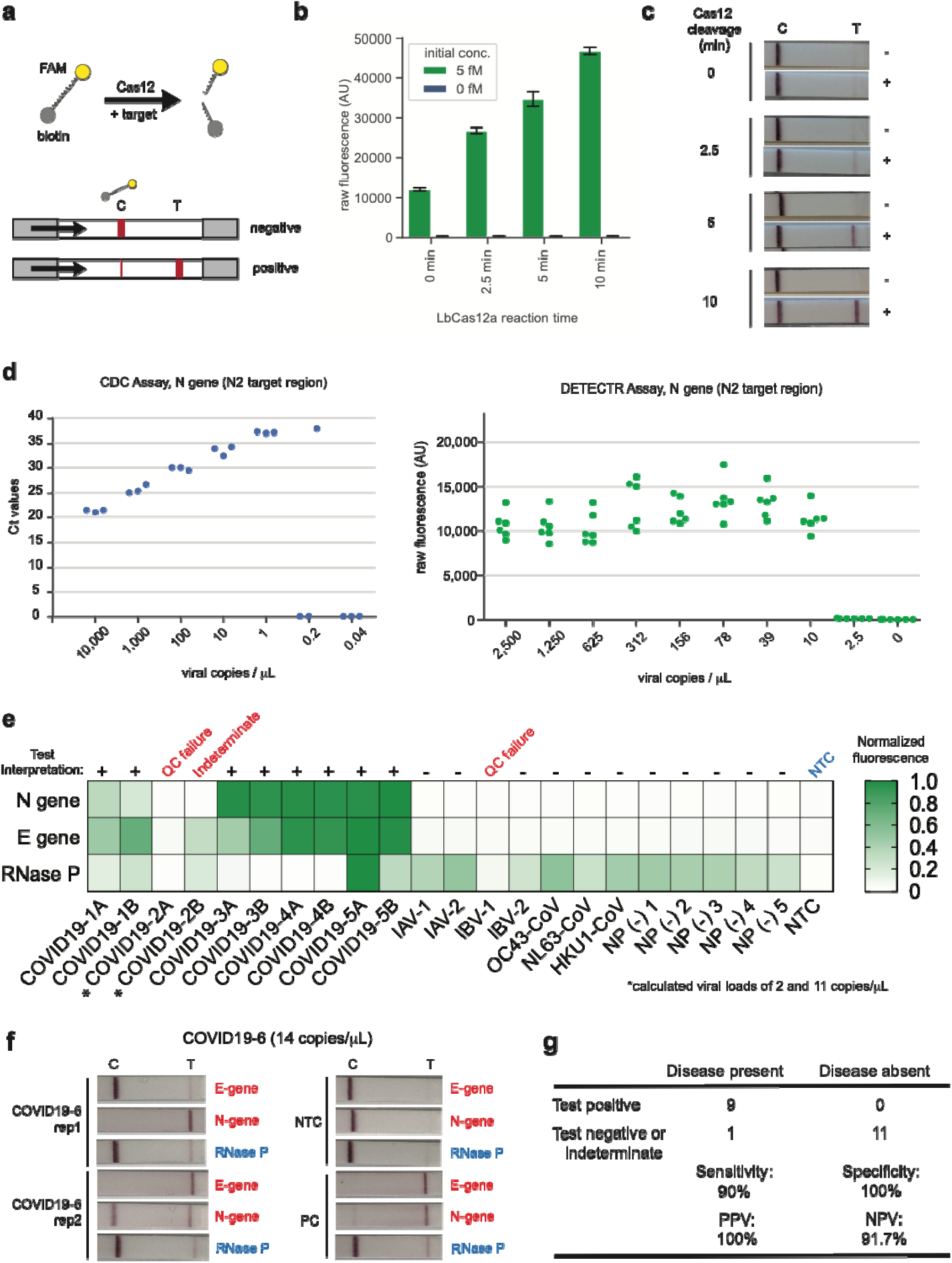
Detection of SARS-CoV-2 in contrived and clinical nasopharyngeal or oropharyngeal swab samples. **(a) Schematic of DETECTR coupled with lateral flow readout**. The intact FAM-biotinylated reporter molecule flows to the control capture line. Upon recognition of the matching target, the Cas-gRNA complex cleaves the reporter molecule, which flows to the target capture line. **(b-c) Comparison of fluorescence to lateral flow. (b)** Fluorescence signal of LbCas12a detection assay on RT-LAMP amplicon for SARS-CoV-2 N-gene saturates within 10 min. RT-LAMP amplicon generated from 2 µL of 5 fM or 0 fM SARS-CoV-2 N-gene IVT RNA by amplifying at 62°C for 20 min. **(c)** LbCas12a on the same RT-LAMP amplicon produces visible signal through lateral flow assay within 5 min. **(d) Limit of detection for CDC qPCR and DETECTR**. Ct values using the CDC qPCR assay (n=3) and fluorescence values using SARS-CoV-2 DETECTR (n=6) using SARS-CoV-2 N2 gene IVT-RNA. **(e) Patient sample DETECTR data**. DETECTR fluorescence values were normalized to the highest value within the N gene, E gene or RNase P set, with a positive threshold at five standard deviations above background. Final determination of the SARS-CoV-2 test was based on the interpretation matrix in Fig. 1e, with results indicated above the heat map. **(f) SARS-CoV-2 DETECTR assay identifies presence of SARS-CoV-2 viral RNA from clinical sample**. Two replicate assays were performed using 2 µL of extracted RNA for each reaction (titer 12 copies/µL). Positive controls used IVT RNA for SARS-CoV-2 targets and total human RNA for RNase P. LbCas12a detection assays were run on lateral flow strips (TwistDx) and imaged after 3 min. **(g) Performance characteristics of the SARS-CoV-2 DETECTR assay**. Abbreviations: fM, femtomolar;NTC, no-template control; PPV, positive predictive value; NPV, negative predictive value.

We then assessed the capability of the RT-LAMP assay to amplify SARS-CoV-2 nucleic acid directly from raw sample matrix consisting of nasopharyngeal swabs from asymptomatic donors placed in universal transport medium (UTM) or phosphate buffered saline (PBS) and spiked with SARS-CoV-2 IVT target RNA. Assay performance was degraded at reaction concentration of ≥10% UTM and ≥20% PBS by volume, with estimated limits of detection decreasing to 500 and 1,500 copies/µL, respectively (**Supplementary Fig. 4)**.

Finally, we tested extracted RNA from 11 respiratory swab samples collected from 6 PCR-positive COVID-19 patients (COVID19-1A/B to COVID19-5A/B, where A=nasopharyngeal swab and B=oropharyngeal swab and COVID19-6, a single nasopharyngeal swab) and 12 nasopharyngeal swab samples from patients with influenza (n=4), common human seasonal coronavirus infections (n=3, representing OC43, HKU1, NL63), and healthy donors (n=5) **(Fig. 2e, f; Supplementary Fig. 5)**. Relative to the CDC qRT-PCR, SARS-CoV-2 DETECTR was 90% sensitive and 100% specific for detection of the coronavirus in respiratory swab samples, corresponding to positive and negative predictive values of 100% and 91.7%, respectively **(Fig. 2g)**.

Here we combined isothermal amplification with CRISPR-Cas12 DETECTR technology to develop a rapid (∼30 min) and low-cost test for detection of SARS-CoV-2 in clinical samples. The use of existing qRT-PCR based assays is hindered by the need for expensive lab instrumentation, and availability is currently restricted to public health laboratories. Importantly, the DETECTR assays developed here have comparable accuracy to qRT-PCR and are broadly accessible, as they use routine protocols and commercially available, “off-the-shelf” reagents. Key advantages of our approach over existing methods such as qRT-PCR include (1) isothermal signal amplification for rapid target detection obviating the need for thermocycling, (2) single nucleotide target specificity (guide RNAs at the N2 site can distinguish SARS-CoV-2 from SARS-CoV and MERS-CoV), (3) integration with portable, low-cost reporting formats such as lateral flow strips, and (4) quick development cycle to address emerging threats from novel zoonotic viruses (<2 weeks for SARS-CoV-2, **Supplementary Fig. 6**).

Although most of the cases of COVID-19 infection during the first month of the epidemic were traced to the city of Wuhan and Hubei province in China, the ongoing rise in cases now appears to be driven to local community transmission^15,16^. For a number of reasons, there is an urgent public health need for rapid diagnostic tests for SARS-CoV-2 infection. The documented cases of asymptomatic infection and transmission in COVID-19 patients^4,5^ greatly expand the pool of individuals who need to be screened. Viral titers in hospitalized patients can fluctuate day-to-day with lack of correlation to disease severity^17-19^, and thus a single negative qRT-PCR test for SARS-CoV-2 does not exclude infection. The virus has also been shown to be shed in stool^20^, raising the possibility of environmental contamination contributing to local disease outbreaks. Low testing platforms such as the DETECTR CRISPR-Cas12 assay developed here may be useful for periodic repeat testing of patient samples. Clinical validation of this assay in response to recent draft guidance from the US FDA^7^ is currently ongoing in a CLIA (Clinical Laboratory Improvement Amendments)-certified microbiology laboratory.

The major pandemics and large-scale epidemics of the past half century have all been caused by zoonotic viruses. Despite these recurrent outbreaks, we still do not have a programmable point of care (POC) diagnostic platform that can be used to promptly address any emerging viral threat. The CRISPR-based DETECTR technology provides such a platform, which we have reconfigured within days to detect SARS-CoV-2 (**Supplementary Fig. 6**). Here we developed a SARS-CoV-2 DETECTR assay, described its performance characteristics and demonstrated compatibility with lateral flow strips. The future development of portable microfluidic-based cartridges to run the assay and use of lyophilized reagents will enable POC testing outside of the clinical diagnostic laboratory, such as airports, local emergency departments and clinics, and other decentralized locations.

## METHODS

### Nucleic acid preparation

SARS-CoV-2 target sequences were designed using all available genomes available from GISAID^21^ as of January 27, 2020. Briefly, viral genomes were aligned using Clustal Omega. Next, LbCas12a target sites on the SARS-CoV-2 genome were filtered against SARS-CoV, two bat-SARS-like-CoV genomes and common human coronavirus genomes. Compatible target sites were finally compared to those used in current protocols from the CDC and WHO. LAMP primers for SARS-CoV-2 were designed against regions of the N-gene and E-gene using PrimerExplorer v5 (https://primerexplorer.jp/e/). RNase P POP7 primers were originally published by Curtis, et al. (2018) and a compatible gRNA was designed to function with these primer sets.

Target RNAs were generated from synthetic gene fragments of the viral genes of interest. First a PCR step was performed on the synthetic gene fragment with a forward primer that contained a T7 promoter. Next, the PCR product was used as the template for an in-vitro transcription (IVT) reaction at 37°C for 2 hours. The IVT reaction was then treated with TURBO DNase (Thermo) for 30 min at 37°C, followed by a heat-denaturation step at 75°C for 15 min. RNA was purified using RNA Clean and Concentrator 5 columns (Zymo Research). RNA was quantified by Nanodrop and Qubit and diluted in nuclease-free water to working concentrations.

### DETECTR assays

DETECTR assays were performed using RT-LAMP for pre-amplification of viral or control RNA targets and LbCas12a for the *trans*-cleavage assay. RT-LAMP was prepared as suggested by New England Biolabs (https://www.neb.com/protocols/2014/10/09/typical-rt-lamp-protocol) with a MgSO_4_ concentration of 6.5 mM and a final volume of 10 µL. LAMP primers were added at a final concentration of 0.2 µM for F3 and B3, 1.6 µM for FIP and BIP, and 0.8 µM for LF and LB. Reactions were performed independently for N-gene, E-gene, and RNase P using 2 µL of input RNA at 62°C for 20 min.

LbCas12a *trans*-cleavage assays were performed similar to those described in Chen, et al. (2018). 50 nM LbCas12a (available from NEB) was pre-incubated with 62.5 nM gRNA in 1X NEBuffer 2.1 for 30 min at 37°C. After formation of the RNA-protein complex, the lateral flow cleavage reporter (/56-FAM/TTATTATT/3Bio/, IDT) was added to the reaction at a final concentration of 500 nM. RNA-protein complexes were used immediately or stored at 4°C for up to 24 hours before use.

### Lateral flow readout

After completion of the pre-amplification step, 2 µL of amplicon was combined with 18 µL of LbCas12a-gRNA complex and 80 µL of 1X NEBuffer 2.1. The 100 µL LbCas12a trans-cleavage assay was allowed to proceed for 10 min at 37°C. A lateral flow strip (Milenia HybriDetect 1, TwistDx) was then added to the reaction tube and a result was visualized after approximately 2 min. A single band, close to the sample application pad indicated a negative result, whereas a single band close to the top of the strip or two bands indicated a positive result.

### Optimized DETECTR method for patient samples

The patient optimized DETECTR assays were performed using RT-LAMP method as described above with the following modifications: A DNA binding dye, SYTO9 (Thermo Fisher Scientific), was included in the reaction to monitor the amplification reaction and the incubation time was extended to 30 min to capture data from lower titre samples.

The fluorescence based patient optimized LbCas12a *trans*-cleavage assays were performed as described above with modifications; 40nM LbCas12a was pre-incubated with 40nM gRNA, after which 100nM of a fluorescent reporter molecule compatible with detection in the presence of the SYTO9 dye (/5Alex594N/TTATTATT/3IAbRQSp/) was added to the complex. 2 µL of amplicon was combined with 18 µL of LbCas12a-gRNA complex in a black 384-well assay plate and monitored for fluorescence using a Tecan plate reader.

### Contrived sample preparation

In-vitro transcribed RNA (gift from California Department of Public Health (CDPH)), with a titer of 10,000 copies/µL (Ct value of 21) was diluted into 2,500 copies/µL first, then serially diluted in water to concentration of 1 250, 625, 312, 156, 78, 39, 10 and 2.5 copies per microliter.

### Human clinical sample collection and preparation

Negative nasopharyngeal swabs were acquired from healthy donors in Chiu lab with the approval of the University of California, San Francisco (UCSF) IRB. Clinical nasopharyngeal and oropharyngeal swab samples of SARS-CoV-2 patients were collected in UTM and transported to the CDPH or UCSF lab. Sample RNA of SARS-CoV-2 was extracted following instructions as described in the CDC EUA-approved protocol^6^ (input 120 µL, elution of 120 µL) using Qiagen DSP Viral RNA Mini Kit (Qiagen) at CDPH and the MagNA Pure 24 instrument (Roche Life Science) at UCSF. Nasopharyngeal swab samples of influenza and common coronavirus were extracted at UCSF using the MagNA Pure 24 instrument.

### CDC real-time qRT-PCR assay

The CDC assay was performed using the ABI 7500 Fast DX instrument (Applied Biosystems) according the CDC EUA-approved protocol^6^.

## Data Availability

The protocol has been released as a white paper online.
Any additional data not in the paper can be provided by the authors upon request.

## SUPPLEMENTARY FIGURES

**Supplementary Figure 1.**
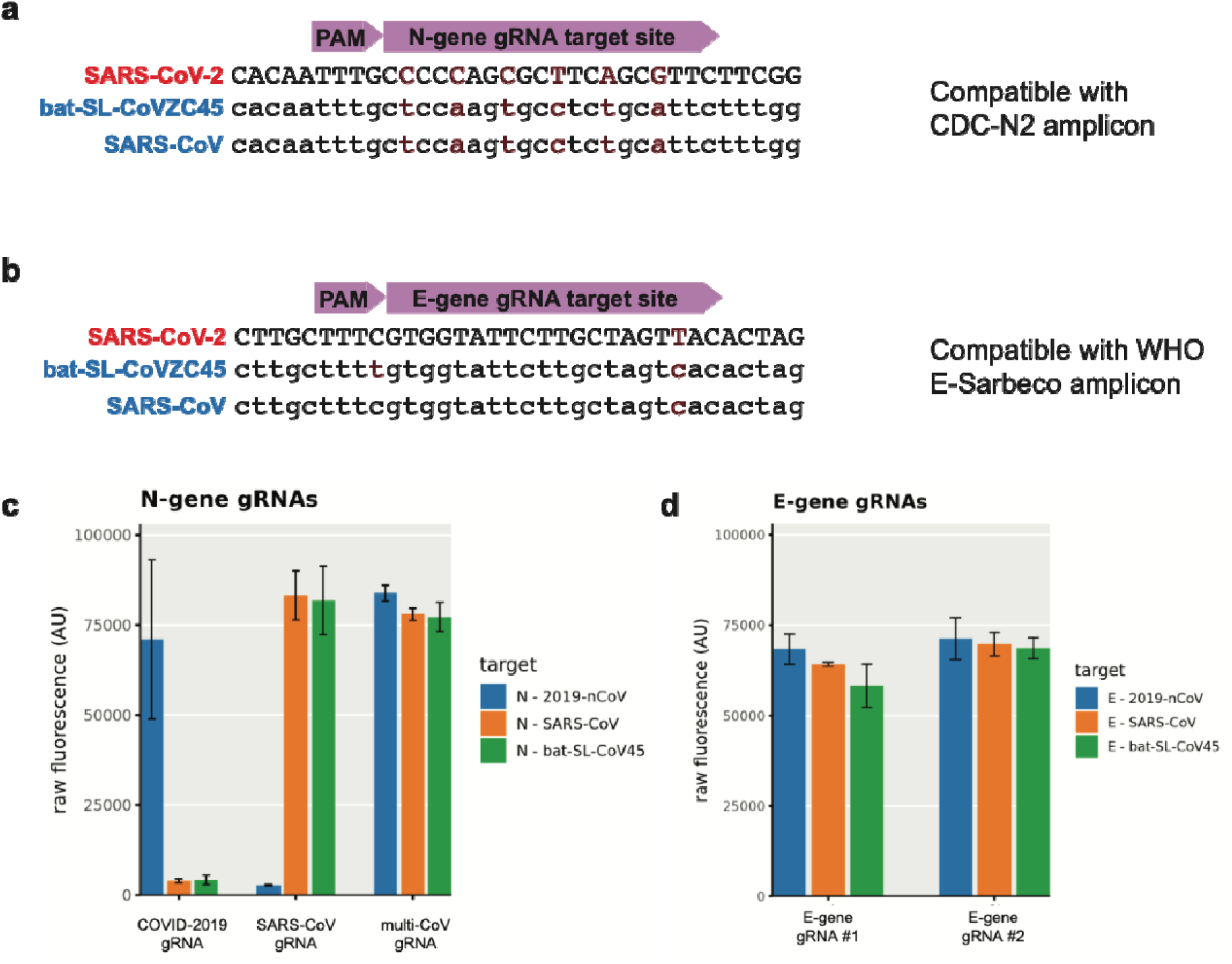
Comparison of sequences between SARS-CoV-2, SARS-CoV, and bat-SL-CoVZC45 at the sites targeted by the gRNAs used in this study. **(a)** The N-gene gRNA is compatible with the CDC-N2 amplicon, and **(b)** the E-gene gRNA is compatible with the WHO E-Sarbeco amplicon. **(c-d)** DETECTR fluorescence values using **(c)** N gene gRNAs and **(d)** E gene gRNAs.

**Supplementary Figure 2.**
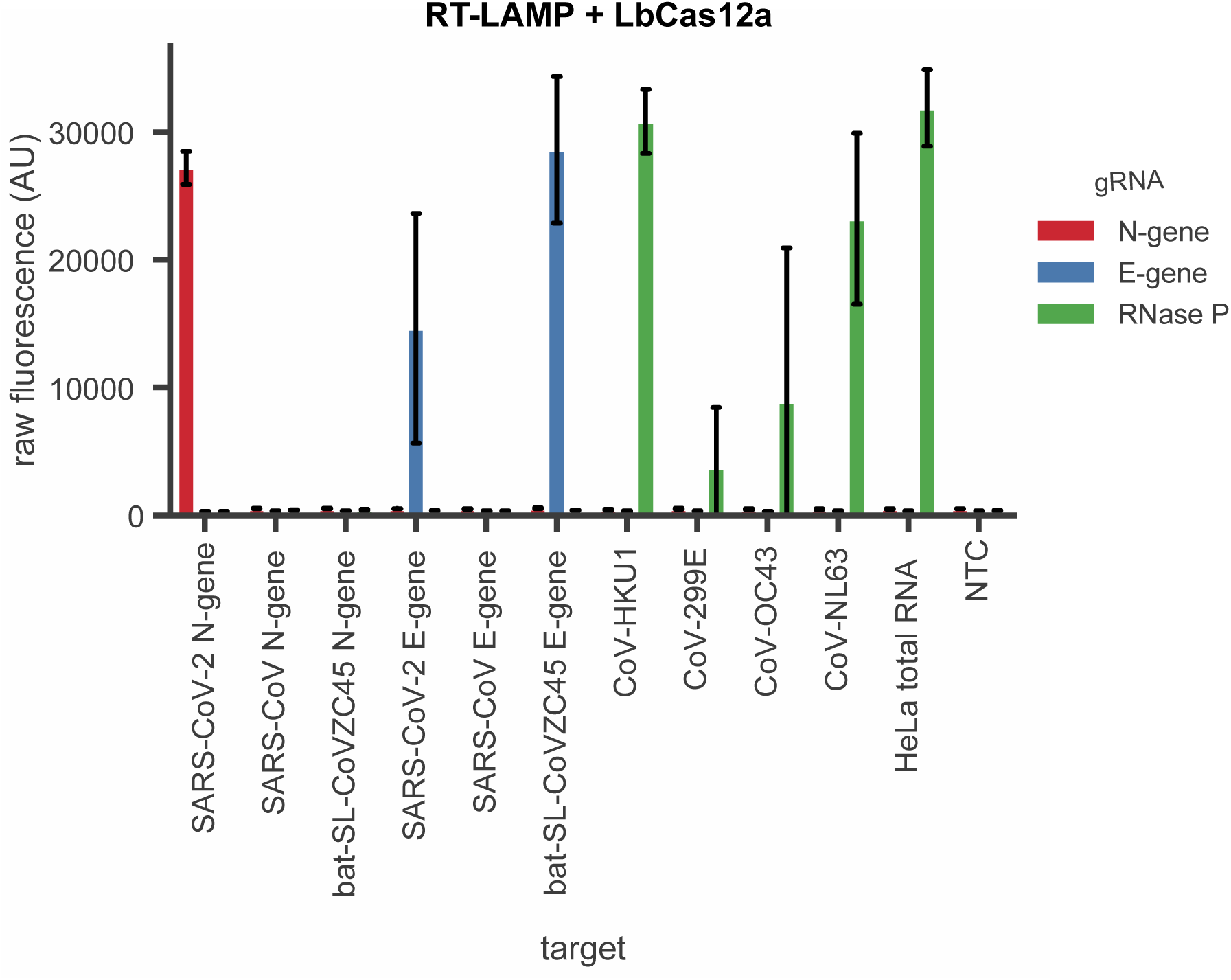
Cross-reactivity of DETECTR to common human coronaviruses. SARS-CoV-2 DETECTR assay (RT-LAMP + Cas12a) was evaluated on IVT RNA products from SARS-CoV-2, SARS-CoV, bast-SL-CoVZC45, and clinical samples from common human coronaviruses. As expected, the N-gene is only detected in SARS-CoV-2, whereas the E-gene is detected only in SARS-CoV-2 and bat-SL-CoVZC45. SARS-CoV E-gene was not detected as the RT-LAMP primer set is not capable of amplifying the SARS-CoV E-gene, even though the E-gene gRNA is capable of detecting the SARS-CoV E-gene target site. RNase P is detected in common human coronaviruses because these samples are RNA extracted from clinical samples. Result shown at 15 min of LbCas12a detection assay signal on fluorescent plate reader.

**Supplementary Figure 3.**
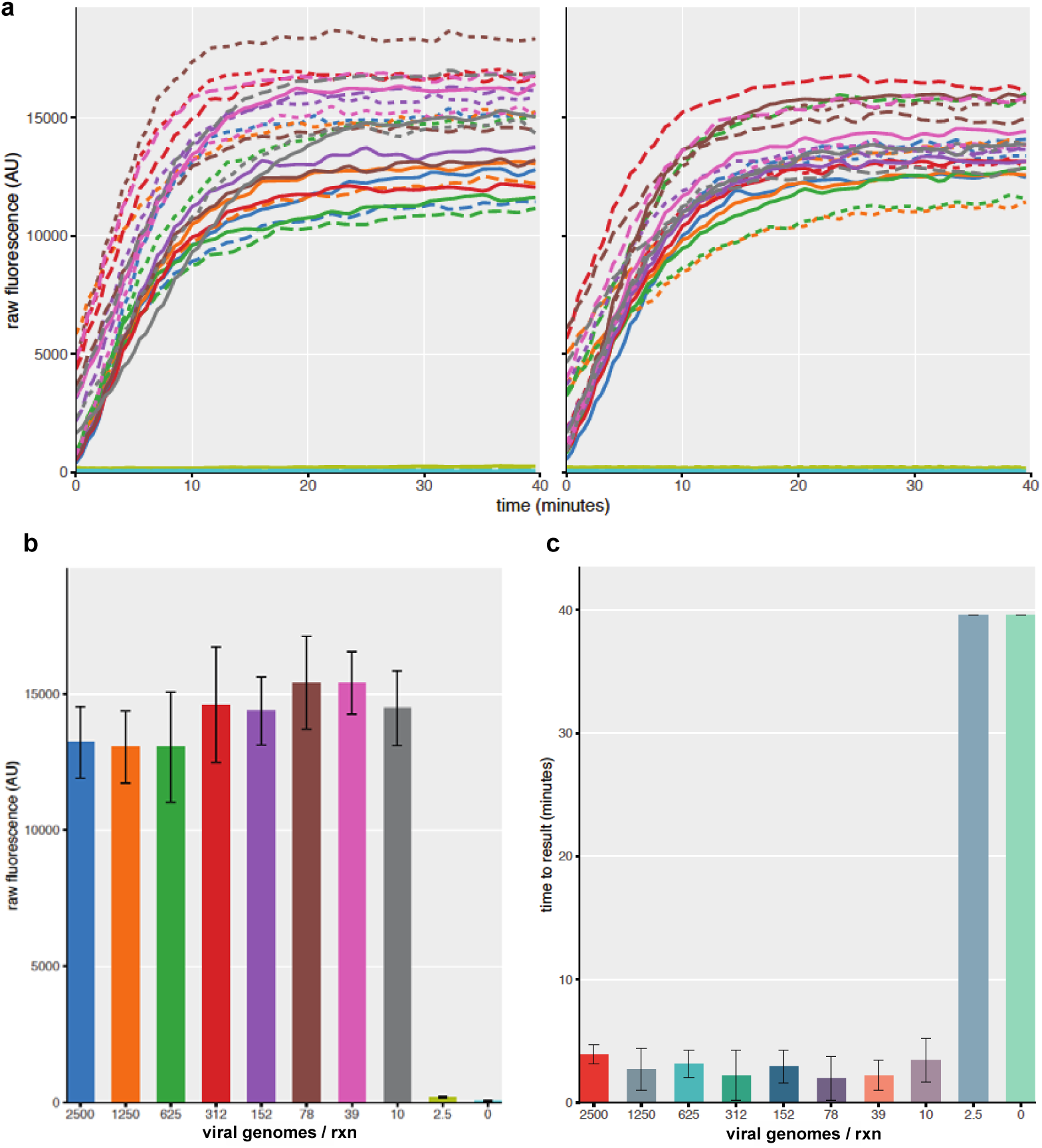
DETECTR analysis of SARS-CoV-2 identifies down to 10 viral genomes in approximately 30 min. Duplicate LAMP reactions were amplified for twenty min followed by LbCas12a DETECTR analysis. **(a)** Raw fluorescence curves generated by LbCas12a detection of SARS-CoV-2 N-gene (n=6) show saturation in less than 20 min. **(b)** Further analysis reveals the limit of detection of the SARS-CoV-2 N-gene to be 10 viral genomes per reaction (n=6). **(c)** Evaluation of the time to result of these reactions highlights detection of 10 viral genomes of SARS-CoV-2 in under 5 min (n=6).

**Supplementary Figure 4.**
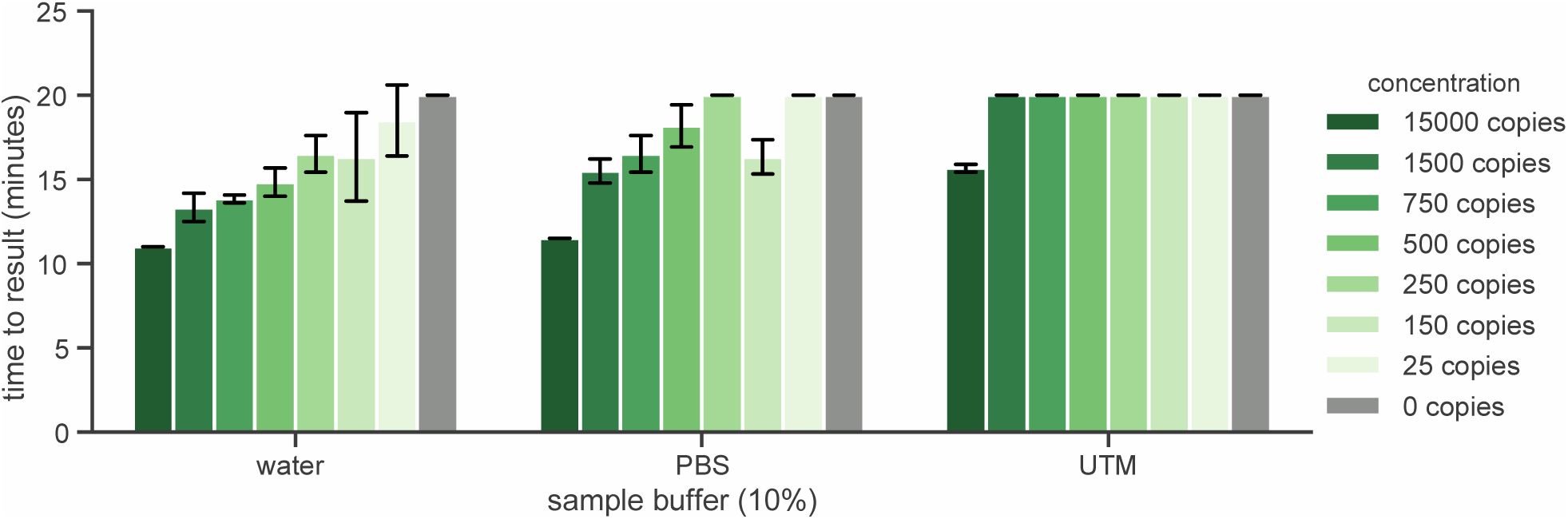
Impact of sample buffers on performance of RT-LAMP pre-amplification. Time-to-result for RT-LAMP amplification (lower value indicates faster amplification) with 10% universal transport medium (UTM), 10% PBS, or 10% water final volume for the SARS-CoV-2 N-gene on a standard curve of the 2019-nCoV positive control plasmid (IDT) in 10% reaction volume. Results indicate that 10% PBS inhibits the assay less than 10% UTM.

**Supplementary Figure 5.**
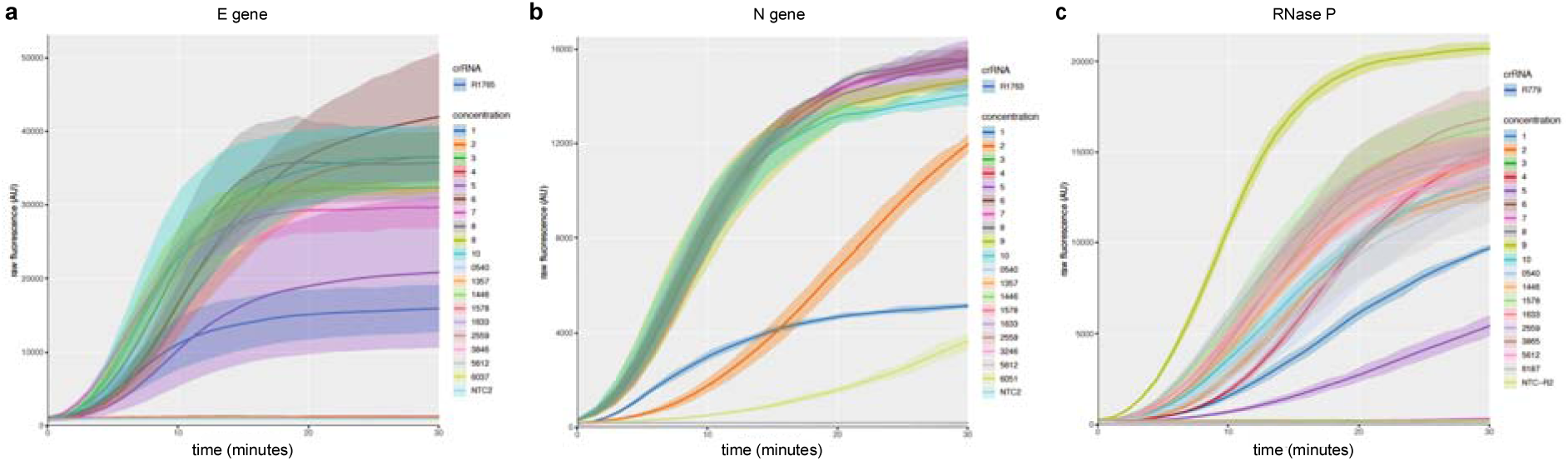
DETECTR kinetic curves on COVID-19 infected patient samples. Ten nasopharyngeal/oropharyngeal swab samples from 5 patients (COVID19-1 to COVID19-5) were tested for SARS-CoV-2 using two different genes, N2 and E as well as a sample input control, RNase P. **(a)** Using the standard amplification and detection conditions, 9 of the 10 patient samples resulted in robust fluorescence curves indicating presence of the SARS-CoV-2 E-gene (20-minute amplification, signal within 10 min). **(b)** The SARS-CoV-2 N-gene required extended amplification time to produce strong fluorescence curves (30-minute amplification, signal within 10 min) for 8 of the 10 patient samples. **(c)** As a sample input control, RNase P was positive for 17 of the 22 total samples tested (20-minute amplification, signal within 10 min).

**Supplementary Figure 6.**
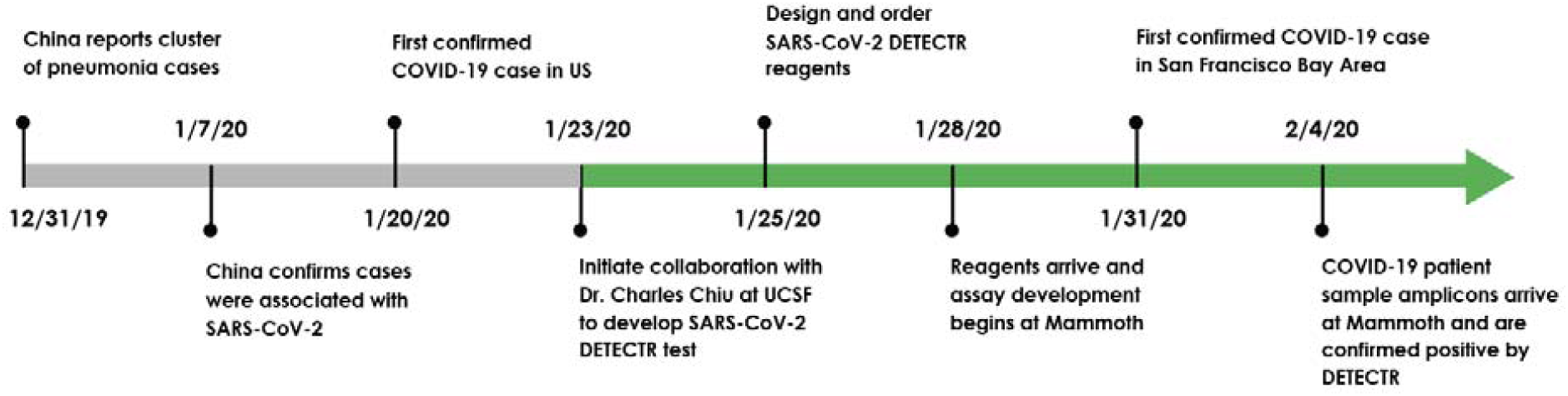
Timeline showing major events in the progression of COVID-19 detection and assay development.

**Extended Data 1 (“Extended_Data_1**.**xlsx”)**. Standard curve generated by running seven 5- or 10-fold dilutions of the CDC N2 qRT-PCR assay, with 3 replicates each dilution. The R-squared measure corresponding to the regression line is 0.9981.

**Extended Data 2 (“Extended_Data_2**.**xlsx”)**. Primer, reporter molecules, target gene fragments, and guide RNAs used in this study.

## ACKNOWLEDGEMENTS

This work was funded by NIH grants R33-AI129455 (CYC) from the National Institute of Allergy and Infectious Diseases and R01-HL105704 (CYC) from the National Heart, Lung, and Blood Institute. We thank Jill Hakim and Dustin Glasner for assisting with sample collection, and Vikram Joshi, Maria-Nefeli Tsaloglou and Xin Miao for helpful discussions in the preparation of this manuscript.

## AUTHOR CONTRIBUTIONS

CYC and JSC conceived the study. JPB conceived, designed and validated DETECTR reagents and protocols. XD and GY validated RT-PCR and LAMP on patient samples. JPB, XD, CLF performed experiments and analyzed data. GY, JS, AG and AS performed experiments. KZ, SM, EH and WG coordinated the study, consented UCSF patients and collected samples. JS, CYP, HG and DW collected samples from patients and extracted the viral RNA. CYC, JPB, XD, and JSC wrote and edited the manuscript. All authors read the manuscript and agree to its contents.

